# Evaluating the determinants of COVID-19 mortality: A cross-country study

**DOI:** 10.1101/2020.05.12.20099093

**Authors:** Jay Squalli

## Abstract

As the COVID-19 pandemic has spread to the entire world, a race to understand the virus and to find an effective and safe vaccine or treatment has triggered interest in the factors contributing to mortality. For instance, some studies have suggested that the BCG vaccine could protect from COVID-19 and nicotine patches could be therapeutic against the virus. This study makes use of data for about 140 countries to evaluate the determinants of COVID-19 mortality. It finds that a country’s share of spending on health care (as a measure of a country’s effectiveness in tracking, recording, and reporting COVID-19 deaths) is positively associated with COVID-19 deaths. It also finds that the share of people above 65 years of age, obesity, and urbanization are all positively associated with COVID-19 mortality. There is no evidence that BCG vaccination, smoking prevalence, and PM25 pollution have any link to COVID-19 mortality. These estimation results are robust to alternative specifications and after controlling for confounding factors and excluding outliers. Policymakers should allocate resources towards the protection of the elderly and those suffering from underlying conditions such as obesity. They should also exercise caution about administering nicotine patches or the BCG vaccine to fight COVID-19 without the backing of concrete scientific evidence.

## 1. Introduction

As the COVID-19 pandemic has spread to the entire world, a race to understand the virus and to find an effective and safe vaccine or treatment has resulted in the emergence of a number of studies interested in the factors that could be contributing to mortality. Among these are reports from the U.S. Centers for Disease Control and Prevention (CDC) providing statistics documenting that a large proportion of people who die from COVID-19 are above 65 years of age (Centers for Disease Control and Prevention, 2020b). Further reports suggest a link between COVID-19 deaths and underlying health conditions, especially obesity (Centers for Disease Control and Prevention, 2020a). Recent research has also posited that countries with current BCG vaccination policy have lower COVID-19-related mortality (Miller et al., 2020), that smoking may have a therapeutic effect on COVID-19 (Farsalinos et al., 2020a,b), and that pollution raises COVID-19 death risk (Wu et al., 2020).

While expedient research about COVID-19 is currently of the utmost importance and urgency, it is crucial that analyses are based on sound statistical research that controls for confounding factors and that does not suffer from biases (e.g. omitted variable bias and self-selection bias). This is particularly problematic in cases where policymakers may rush to unnecessarily recommend the use of nicotine patches as a potential treatment against COVID-19 despite limited evidence (Farsalinos et al., 2020b). Indeed, the research claiming that the BCG vaccine protects against COVID-19 has already triggered randomized controlled trials in the Netherlands and Australia (Curtis et al., 2020). Given the seriousness of the pandemic and the importance of understanding the virus, the present study aims to evaluate the determinants of COVID-19 mortality including but not limited to the above hypothesized factors. To this end, it is organized as follows: Section 2 describes the data and methodology. Section 3 summarizes the results. Section 4 assesses robustness. Section 5 concludes.

## 2. Data and Methodology

Data on COVID-19 mortality and other relevant variables were extracted from worldometers.info on May 4, 2020. All data are from 2018 or the latest available year. Data for population, health care spending (% of GDP), share of population above 65 years, urban population (% of total population), smoking prevalence (% of men and women 15+), and particulate matter 2.5 mean annual exposure *(μg* per *m*^3^) are from the World Bank. BCG data are from the BCG World Atlas. Obesity (% of adults with BMI ≥ 30) data are from the World Health Organization. After combining all variables, the sample is comprised of more than 140 countries.

The following represents a specification based on our current understanding of the COVID-19 virus and related hypotheses:

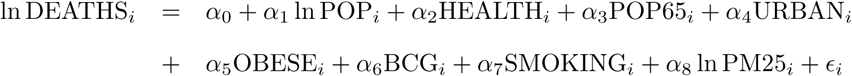

where the dependent variable represents the number of reported COVID-19 deaths for country i. This is estimated as a function of total population (POP) as a scale variable, health care spending as % of GDP (HEALTH), the share of the population above 65 years of age (POP65), urban population (% of the total population), the obesity rate (% of adults with BMI ≥ 30), a dummy variable taking a value of one when a country has a BCG vaccination policy and zero otherwise (BCG), smoking prevalence (% of men and women 15+), and particulate matter 2.5 mean annual exposure (*μg* per *m*^3^).

Since the dependent variable is measured in levels, it is important to introduce population as a scale variable. The HEALTH variable is introduced to control for the provision of health care across the world. The variable POP65 is introduced as an explanatory variable in line with the CDC’s contention that a large proportion of people dying from COVID-19 are above the age of 65 (Centers for Disease Control and Prevention, 2020b). The variable URBAN is introduced to control for urban areas’ role in spreading outbreaks (Whiting, 2020). The OBESE variable is introduced to account for the CDC’s observation that a large share of deaths are patients that suffer from underlying conditions, especially obesity (Centers for Disease Control and Prevention, 2020a). The BCG dummy variable is introduced to verify the contention that countries with BCG vaccination policies suffer fewer COVID-19 deaths (Miller et al., 2020). The SMOKING variable is also introduced to verify the claims that smoking could have a therapeutic role against COVID-19 and could protect people from contracting the virus (Farsalinos et al., 2020b,a). Finally, the PM25 variable is introduced to account for the contention that pollution exacerbates COVID-19 symptoms and increases the death risk from the virus (Wu et al., 2020).

All estimations are completed using a Least Squares estimator with bootstrapped standard errors. This estimator is particularly useful because it allays concerns about within-sample distortions and derives estimates of standard errors and confidence intervals based on the underlying distribution of the sample rather than based on some a priori distributional assumptions. All estimations are completed with 500 bootstrap replications and are expressed in log-linear form in order to interpret the coefficient estimates as elasticities.

## 3. Results

Estimation results are summarized in Table 1. The base model in column (1) shows that the coefficient estimate for the share of spending on health care is positive and statistically significant (*p <* 0.001). The introduction of the share of the population 65+ variable in column (2) causes the coefficient estimate for the population variable to approach unity and that for the health care variable to decrease in magnitude and in statistical significance (*p* < 0.01). The coefficient estimate for the share of the population 65+ is positive and statistically significant (*p* < 0.001). Column (3) shows estimation results of a model augmented with the obesity rate variable. The latter’s coefficient estimate is positive and statistically significant (*p* < 0.001). The coefficient estimate for the health care variable decreases again in magnitude and statistical significance (*p* < 0.05), while the coefficient estimate for the share of the population 65+ decreases slightly in magnitude but remains at the same level of statistical significance. The introduction of the urban population variable in column (4) reduces the magnitude and statistical significance of the coefficient estimate for the obesity rate (*p <* 0.01) and only marginally affects the rest. The coefficient estimate for the urban population variable is not statistically significant. In column (5), the introduction of the BCG vaccination variable has virtually no effect on the estimation results. In fact, prior to its introduction, about 74.7% of the variation in COVID-19 deaths was explained by the explanatory variables in column (4). This percentage remains the same after the introduction of the BCG vaccination variable, suggesting that the latter variable yields no additional explanatory power. Columns (6) and (7) report estimation results that introduce the smoking prevalence and PM25 variables, respectively. The coefficient estimates for both variables are not statistically significant and the corresponding adjusted R^2^ decline to 0.745 and 0.742, respectively. In addition, the fact that the coefficient estimate for health spending loses statistical significance in column (7) suggests that there exists some interaction effect with the urban population (as a measure of urbanization), smoking prevalence, and PM25 pollution. Intuitively, this interaction could be explained by how urbanization, particulate matter pollution, and smoking prevalence could represent a significant trigger of related health care spending. However, the absence of statistical significance and lower adjusted R^2^ values suggest that these two variables, just like the BCG vaccination variable, have no explanatory power.

**Table 1:** Estimation results

|  | (1) | (2) | (3) | (4) | (5) | (6) | (7) |
| --- | --- | --- | --- | --- | --- | --- | --- |
| ln population | 0.776***<br>(0.0821) | 0.904***<br>(0.0680) | 0.977***<br>(0.0604) | 0.974***<br>(0.0627) | 0.975***<br>(0.0651) | 1.057***<br>(0.0691) | 1.065***<br>(0.0736) |
| Health spending | 0.401***<br>(0.0625) | 0.148**<br>(0.0516) | 0.107*<br>(0.0476) | 0.115*<br>(0.0486) | 0.117*<br>(0.0495) | 0.113*<br>(0.0473) | 0.107<br>(0.0575) |
| Share of<br>population 65+ |  | 0.188***<br>(0.0222) | 0.165***<br>(0.0205) | 0.155***<br>(0.0217) | 0.152***<br>(0.0210) | 0.140***<br>(0.0264) | 0.135***<br>(0.0302) |
| Obesity rate |  |  | 0.0720***<br>(0.0141) | 0.0547**<br>(0.0177) | 0.0548**<br>(0.0169) | 0.0554**<br>(0.0196) | 0.0562**<br>(0.0212) |
| Urban population |  |  |  | 0.0123<br>(0.00728) | 0.0130<br>(0.00731) | 0.0200*<br>(0.00836) | 0.0196*<br>(0.00871) |
| BCG vaccination |  |  |  |  | 0.220<br>(0.226) | 0.314<br>(0.252) | 0.306<br>(0.252) |
| Smoking prevalence |  |  |  |  |  | -0.00567<br>(0.0163) | -0.00463<br>(0.0158) |
| ln PM25 |  |  |  |  |  |  | -0.0921<br>(0.280) |
| Intercept | -11.13***<br>(1.409) | -13.39***<br>(1.160) | -15.44***<br>(1.032) | -15.80***<br>(1.142) | -15.96***<br>(1.175) | -17.52***<br>(1.246) | -17.29***<br>(1.424) |
| adj. $R^2$ | 0.502 | 0.697 | 0.743 | 0.747 | 0.747 | 0.745 | 0.742 |
| $N$ | 145 | 142 | 140 | 140 | 140 | 117 | 117 |

Bootstrap standard errors in parentheses
\* $p < 0.05$ , \*\* $p < 0.01$ , \*\*\* $p < 0.001$

In sum, focusing on the estimation results in column (4), which yield the best goodness-of-fit, the evidence suggests that countries that have a larger share of health spending have greater COVID-19 mortality. While a high spending share on health care could suggest a large health care sector, it is not indicative of quality of health care. For instance, based on World Bank data, the United States spent more than 17% of its GDP on health care (second to Vanuatu) in 2017 and managed to get the world’s highest number of COVID-19 deaths in absolute terms and the ninth largest in per capita terms (after excluding island nations). In fact, most European countries that have relatively high spending on health care also happen to have a high number of deaths in absolute and in capita terms. Another important observation is that a number of countries that have low health care spending have reported a relatively low number of deaths. In such countries, extreme lock-down and confinement measures would have been warranted after the realization that the pandemic would be too overwhelming and could lead to greater mortality and a collapse of the health care system. Of course, fewer reported deaths could also be the result of insufficient resources to track and report all deaths related to COVID-19.

The evidence also suggests that countries that have a larger share of the population above 65 years of age have greater COVID-19 mortality, consistent with Centers for Disease Control and Prevention (2020b). Countries with a higher obesity rate also have greater COVID-19 mortality, consistent with Centers for Disease Control and Prevention (2020a). There is no evidence suggesting a link between BCG vaccination and COVID-19 deaths, contrary to Miller et al. (2020). There also exists no link between smoking prevalence or pollution on COVID-19 mortality, contrary to Farsalinos et al. (2020b,a) and Wu et al. (2020), respectively.

## 4. Robustness of the Results

There are always concerns about potentially influential observations when using cross-country studies. This can be addressed by plotting leverages against normalized squared residuals. Leverages capture the influence of observed values on fitted values and represent the diagonal elements of the hat matrix *(h_ii_)* and are bounded by the limits 1/*n* and 1. An observation is considered a leverage when *h_ii_ >* (2*k* + 2)/*n*, where *k* represents the number of coefficients (Welsch & Kuh, 1977). To minimize the number of explanatory variables used to explain variation in COVID-19 mortality and to increase robustness, the estimated specification is simplified by parsimony through the elimination of variables that are statistically insignificant and have large p values. Based on the previous estimations, the model is reduced to exclude the SMOKING and PM25 variables. This reduced model is used as a base model to derive leverage estimates. Given a sample of 140 countries and 7 coefficients, this would suggest that a particular country would be considered an outlier if its leverage point *h_ii_ >* 0.114. Estimates of leverage values for all countries, as shown in Figure 1, indicate that there are four countries with leverage values exceeding this level, namely the United States (0.185), Japan (0.175), Sierra Leone (0.139), and Singapore (0.124).

**Figure 1:**
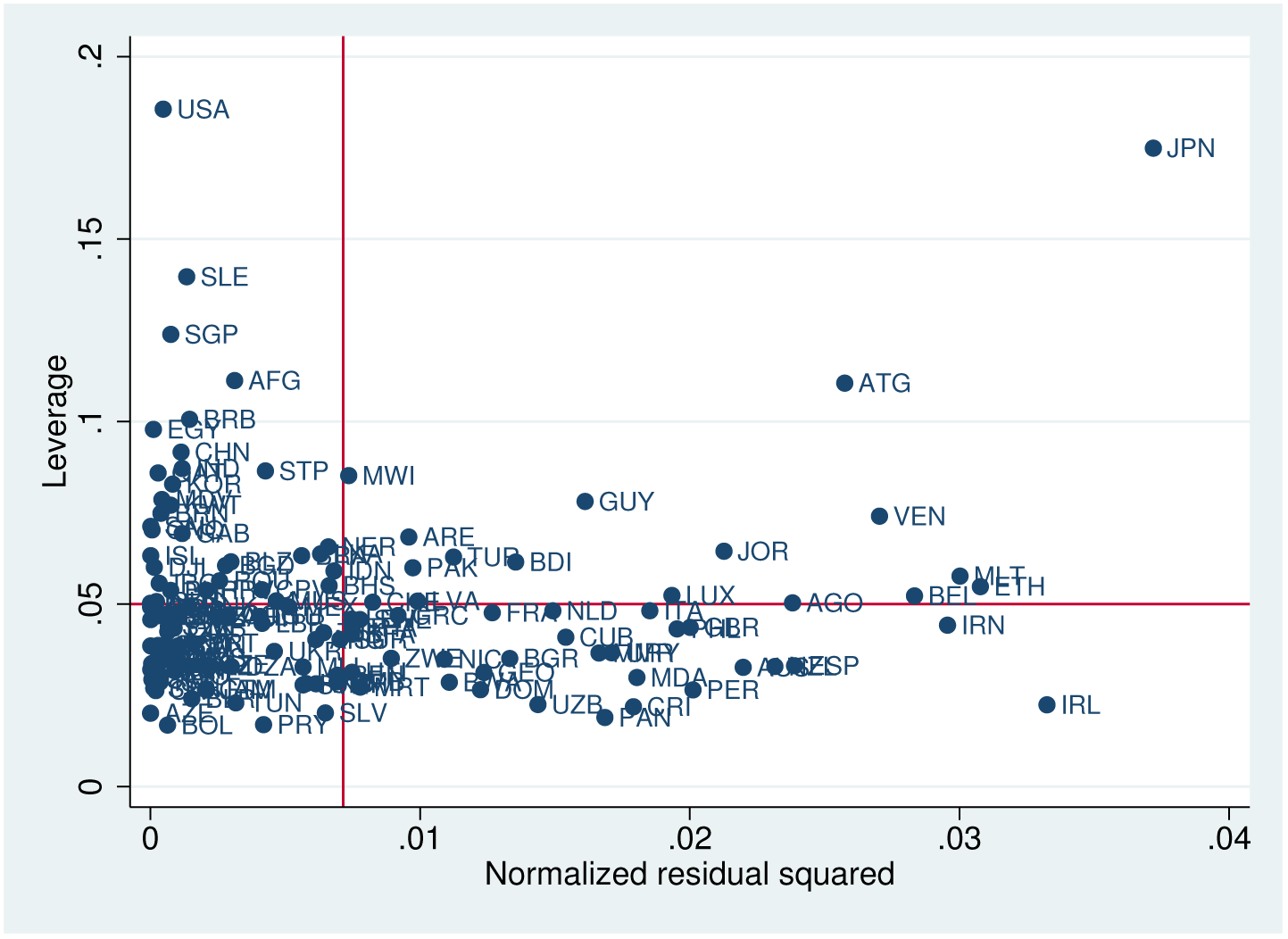
Plot of leverage and normalized residual squared values for the detection of influential observations

Table 2 summarizes estimation results after the exclusion of these countries. The key new finding is that the coefficient estimate for the urban population variable is positive and statistically significant *(p <* 0.05) in the estimation results reported in column (4), which yields the highest adjusted R^2^.^1^ This suggests that urbanization may have played a significant role in the spreading of COVID-19 and ensuing deaths. It is worth noting that this is likely related to the spreading of diseases within dense populations in urban areas rather than to pollution created by cities. In fact, this is confirmed by the lack of statistical significance for the coefficient estimate of PM25.

**Table 2:** Estimation results without outliers

|  | (1) | (2) | (3) | (4) | (5) | (6) | (7) |
| --- | --- | --- | --- | --- | --- | --- | --- |
| ln population | 0.784***<br>(0.0851) | 0.917***<br>(0.0637) | 0.986***<br>(0.0656) | 0.981***<br>(0.0645) | 0.982***<br>(0.0687) | 1.072***<br>(0.0734) | 1.070***<br>(0.0714) |
| Health spending | 0.443***<br>(0.0651) | 0.147*<br>(0.0616) | 0.115*<br>(0.0572) | 0.124*<br>(0.0582) | 0.127*<br>(0.0570) | 0.127*<br>(0.0610) | 0.128<br>(0.0685) |
| Share of<br>population 65+ |  | 0.198***<br>(0.0222) | 0.175***<br>(0.0209) | 0.164***<br>(0.0206) | 0.161***<br>(0.0215) | 0.151***<br>(0.0260) | 0.152***<br>(0.0323) |
| Obesity rate |  |  | 0.0643***<br>(0.0138) | 0.0361*<br>(0.0166) | 0.0364*<br>(0.0185) | 0.0309<br>(0.0194) | 0.0305<br>(0.0206) |
| Urban population |  |  |  | 0.0175*<br>(0.00735) | 0.0180*<br>(0.00825) | 0.0279**<br>(0.00897) | 0.0280**<br>(0.00898) |
| BCG vaccination |  |  |  |  | 0.163<br>(0.229) | 0.266<br>(0.235) | 0.267<br>(0.240) |
| Smoking prevalence |  |  |  |  |  | -0.00787<br>(0.0165) | -0.00812<br>(0.0167) |
| ln PM25 |  |  |  |  |  |  | 0.0217<br>(0.268) |
| Intercept | -11.50***<br>(1.527) | -13.66***<br>(1.106) | -15.56***<br>(1.181) | -16.00***<br>(1.226) | -16.13***<br>(1.269) | -17.88***<br>(1.377) | -17.93***<br>(1.415) |
| adj. $R^2$ | 0.495 | 0.701 | 0.735 | 0.744 | 0.743 | 0.744 | 0.742 |
| $N$ | 141 | 138 | 136 | 136 | 136 | 113 | 113 |

Bootstrap standard errors in parentheses
\* $p < 0.05$ , \*\* $p < 0.01$ , \*\*\* $p < 0.001$

In sum, despite the exclusion of the United States, as the country with the largest number of COVID-19 deaths, and other outliers, the overall conclusions advanced earlier still hold. That is, countries with a greater share of spending on healthcare, a larger proportion of people above 65 years of age, and a larger obesity rate have greater COVID-19 mortality. When interpreted as elasticities, these results suggest that a 10% in spending on health care is associated with a 1.24% greater reported COVID-19 mortality. Individual increases of 10% in obesity and in the urban population are also associated with corresponding increases of 0.36% and 0.17% on COVID-19 deaths, respectively. Finally, and contrary to previous research, there is no evidence of a link between COVID-19 mortality and BCG vaccination, smoking prevalence, and PM25 pollution.

## 5. Discussion and Conclusions

This paper evaluates the determinants of COVID-19 mortality using a sample of 140 countries. It presents seven key findings. First, countries that have a larger share of health care spending have greater (reported) COVID-19 mortality. It is this paper’s contention that greater spending likely reflects a country’s ability to better track, document, and report COVID-19 deaths. This suggests that countries with low spending on health care may not have the resources to accurately report COVID-19 deaths and may even have an incentive to underreport. Second, consistent with Centers for Disease Control and Prevention (2020b), countries with a greater share of the population older than 65 years of age have greater COVID-19 mortality. Third, countries that suffer from a high obesity rate have greater COVID-19 mortality, consistent with Centers for Disease Control and Prevention (2020a). Fourth, highly urbanized countries have greater COVID-19 mortality, consistent with the contention that the COVID-19 pandemic may have spread faster in urbanized areas (Whiting, 2020). Fifth, there is no evidence of a link between BCG vaccination and COVID-19 mortality. Sixth, there is also no evidence of a link between smoking prevalence and COVID-19 deaths. Finally, there is no evidence of an association between PM25 pollution and COVID-19 deaths.

There are evidently other factors not included in these estimations that could contribute to COVID-19 mortality. For instance, a country’s timely response to the pandemic, irrespective of its spending on health care, could have measurable effects on COVID-19 mortality. New Zealand is an important success story (Cousins, 2020), whereas the United States, to this day, has had one of the worst responses in the world (Blodget & Plotz, 2020). Countries that have implemented and enforced extreme lock-down measures have likely fared better in terms of COVID-19 deaths than those that did not.

This research has important policy implications. Policymakers should allocate resources towards the protection of the most vulnerable members of society from the pandemic, namely the elderly and those suffering from underlying conditions such as obesity. They should also exercise caution about the potential dangers of promoting certain unvetted drugs, implementing randomized controlled trials, or administering nicotine patches or the BCG vaccine without the backing of concrete scientific evidence.

## Data Availability

All data are available for free in the public domain.

## Footnotes

1 Although the estimations in column (6) also yield an adjusted R^2^ of 0.744, those in column (4) are preferable due to the larger sample.

